# Social and Clinical Determinants of COVID-19 Outcomes: Modeling Real-World Data from a Pandemic Epicenter

**DOI:** 10.1101/2021.04.06.21254728

**Authors:** Jyothi Manohar, Sajjad Abedian, Rachel Martini, Scott Kulm, Mirella Salvatore, Kaylee Ho, Paul Christos, Thomas Campion, Julianne Imperato-McGinley, Said Ibrahim, Teresa H. Evering, Erica Phillips, Rulla Tamimi, Vivian Bea, Onyinye D. Balogun, Andrea Sboner, Olivier Elemento, Melissa Boneta Davis

## Abstract

**IMPORTANCE:** As the United States continues to accumulate COVID-19 cases and deaths, and disparities persist, defining the impact of risk factors for poor outcomes across patient groups is imperative.

**OBJECTIVE:** Our objective is to use real-world healthcare data to quantify the impact of demographic, clinical, and social determinants associated with adverse COVID-19 outcomes, to identify high-risk scenarios and dynamics of risk among racial and ethnic groups.

**DESIGN:** A retrospective cohort of COVID-19 patients diagnosed between March 1 and August 20, 2020. Fully adjusted logistical regression models for hospitalization, severe disease and mortality outcomes across 1-the entire cohort and 2-within self-reported race/ethnicity groups.

**SETTING:** Three sites of the NewYork-Presbyterian health care system serving all boroughs of New York City. Data was obtained through automated data abstraction from electronic medical records.

**PARTICIPANTS:** During the study timeframe, 110,498 individuals were tested for SARS-CoV-2 in the NewYork-Presbyterian health care system; 11,930 patients were confirmed for COVID-19 by RT-PCR or covid-19 clinical diagnosis.

**MAIN OUTCOMES AND MEASURES:** The predictors of interest were patient race/ethnicity, and covariates included demographics, comorbidities, and census tract neighborhood socio-economic status. The outcomes of interest were COVID-19 hospitalization, severe disease, and death.

**RESULTS:** Of confirmed COVID-19 patients, 4,895 were hospitalized, 1,070 developed severe disease and 1,654 suffered COVID-19 related death. Clinical factors had stronger impacts than social determinants and several showed race-group specificities, which varied among outcomes. The most significant factors in our all-patients models included: age over 80 (OR=5.78, p= 2.29×10^−24^) and hypertension (OR=1.89, p=1.26×10^−10^) having the highest impact on hospitalization, while Type 2 Diabetes was associated with all three outcomes (hospitalization: OR=1.48, p=1.39×10^−04^; severe disease: OR=1.46, p=4.47×10^−09^; mortality: OR=1.27, p=0.001). In race-specific models, COPD increased risk of hospitalization only in Non-Hispanics (NH)-Whites (OR=2.70, p=0.009). Obesity (BMI 30+) showed race-specific risk with severe disease NH-Whites (OR=1.48, p=0.038) and NH-Blacks (OR=1.77, p=0.025). For mortality, Cancer was the only risk factor in Hispanics (OR=1.97, p=0.043), and heart failure was only a risk in NH-Asians (OR=2.62, p=0.001).

**CONCLUSIONS AND RELEVANCE:** Comorbidities were more influential on COVID-19 outcomes than social determinants, suggesting clinical factors are more predictive of adverse trajectory than social factors.

**KEY POINTS:** *QUESTION:* What is the impact of patient self-reported race, ethnicity, socioeconomic status, and clinical profile on COVID-19 hospitalizations, severity, and mortality?

*FINDINGS:* In patients diagnosed with COVID-19, being over 50 years of age, having type 2 diabetes and hypertension were the most important risk factors for hospitalization and severe outcomes regardless of patient race or socioeconomic status.

*MEANING:* In this large sample pf patients diagnosed with COVID-19 in New York City, we found that clinical comorbidity, more so than social determinants of health, was associated with important patient outcomes.

## INTRODUCTION

As of March 1, 2021, over 25 million coronavirus disease 2019 (COVID-19) cases have resulted in more than 500,000 deaths in the US. New York City (NYC) bore the brunt of the initial wave of infections with over 60,000 hospitalized cases and 10,000 deaths. Early in the crisis, it was clear that the incidence and outcome of COVID-19 infection differed vastly across patient populations. For example, older, male individuals with clinical co-morbidities were shown to have worst outcomes than other groups^1^. Ethnic and racial disparities in COVID-19 incidence and mortality also emerged in the US, where African American and Hispanic groups suffer disproportionate incidence^1-5^ and related death rates^3,5^. These disparities may partially be explained by the prevalence of comorbidities^5,6^. However, recent investigations also suggest factors related to socioeconomic status, such as employment and neighborhood density may play a role. Specifically, individuals living in higher poverty and/or higher density areas were found to have higher COVID-19 infection rates, and residents in COVID-19 hotspot areas were more likely to be younger, non-White, and have multiple co-morbidities, compared to those who live in areas with lower risk of COVID-19 infection^2,3,5^. The influence of poverty levels on death outcomes are less clear, as some groups have reported higher death rates among individuals living in higher poverty areas^3^, and others have reported a protective survival benefit among those in high poverty^5^ areas or with non-White race/ethnicity^6^.

Despite many studies, a full understanding of specific factors of differential risk of disease trajectory across diverse populations is still unclear. In particular, the role of social determinants of health (SDOH) when controlled for clinical factors remains unclear. Most importantly, risk analyses have only been performed in the general population and variance of risk factors among racial/ethnic group is unknown. Finally, whether risk factor status can be developed into predictive outcome models to be used in high-risk patient triage to prevent severe outcomes is equally unknown.

To address these issues, we used a real-world dataset (RWD) from NewYork-Presbyterian (NYP) Hospitals, a healthcare system that provides primary and specialty clinical services for a diverse population of patients from all five NYC boroughs, and was a major contributor to disease management during the initial COVID-19 spike in NYC. Specifically, we sought to quantify the relative impact of risk factors related to patient demographics, social determinants of health and clinical comorbidities on COVID-19 disease outcomes across patient groups. We hypothesized that multivariate modeling in a large and diverse cohort would reveal the contribution of each factor and help identify specific reasons why certain population groups have had worse outcomes, compared to others. We anticipate these findings will facilitate better triage and prioritization of patients with high-risk of disease severity and/or mortality.

## METHODS

### Setting

Weill Cornell Medicine (WCM), located in New York City, has over 20 outpatient sites across the city. WCM physicians also hold admitting privileges at NewYork-Presbyterian (NYP) Hospital. WCM is an academic medical center, approximately 1,000 attending physicians and over 250,000 patient visits per year. This study was approved by the WCM Institutional Review Board (IRB).

### Data Sources *- Integrated Data Registry/Repository (IDR)*

Our system clinicians used the EpicCare^®^ Ambulatory Electronic Health Record (EHR) system in conjunction with AllScripts Acute EHR system to document clinical care in the outpatient and inpatient settings, respectively. We aggregated raw data from all these disparate EHR source systems which then were transformed and modeled in WCM’s COVID centralized data repository, COVID Institutional Data Repository (IDR)^7^. The NYP hospital system catchment includes patients from all NYC boroughs. Of the 100,000+ patients in our COVID IDR, which represented three NYP sites – designated as Sites 1, 2 and 3, we included patients with confirmed COVID-19.

### Participants

COVID-19 confirmed cases were defined as patients that had nasopharyngeal swab PCR testing performed with “Detected” results or those who received a COVID-19 ICD-10 diagnosis (excluding those that were also confirmed as “Not Detected” by PCR assay). We utilized self-reported race and ethnicity subcategories that were recorded as separate fields in the EHR. Patients who identified as “Hispanic or Latino or Spanish Origin” ethnicity were designated as “Hispanic or Latino or Spanish Origin”, and all others were classified as the Non-Hispanic race categories they identified. Thus, we designated patients to mutually exclusive self-identified categories: Hispanic (HISP); Non-Hispanic Black (NH-Black); Non-Hispanic White (NH-White); and Non-Hispanic Asian (NH-Asian) (Table 1.)

For the race-specific regression analyses, age was binned as either less than 50 years or 50+ years. We extracted co-morbidity data using ICD-10 Codes as documented in the EHR (ICD10 codes: type 2 diabetes (T2D), E11; type 1 diabetes (T1D), E10; hypertension (HTN), I10; heart failure (HF), I50; cardiovascular disease (CVD), I25; cancer, C80; chronic obstructive pulmonary disease (COPD), J44; asthma, J45; depression, F32). Body Mass Index (BMI) within three months of COVID-19 diagnosis was extracted from the EHR. This variable was then categorized as “<30 (non-obese)” or “30+ (obese)”. Hospital Site was defined as the location at which the patient was either tested for COVID-19 or first received a COVID-19 ICD-10 diagnosis. Social Determinants of Health (SDOH) were quantified using the Neighborhood Deprivation Index (NDI), calculated as previously described in Messer et al^8^ using census tract data summarizing five socio-demographic domains associated with health outcomes, including income/poverty, education, employment, housing, and occupation, using principal components analysis. NDI was assigned to each patient using the longitude and latitude coordinates of street addresses, and this value was categorized into three ranges: [-2.07, -0.696] (low), (−0.696, 0.544] (medium), and (0.544, 3.09] (high). We classified available insurance data for patients as either Commercial, Medicare, Medicaid or Hybrid (i.e., different primary and secondary insurances). The severe disease category included hospitalized patients who were intubated, were on vasopressors, had a diagnosis of acute respiratory distress syndrome (ARDS), and/or were on kidney dialysis (ICD10 codes: ARDS, J80; renal dialysis, Z99.2 or Z49). These criteria were based on criteria from the WHO Ordinal Scale for Clinical Improvement, as well as clinical manifestation of severe disease in COVID-19 patients^9^.

### Statistical Analysis

Demographic, clinical, and socioeconomic characteristics of our cohort were summarized using descriptive statistics for each model group; including COVID-19 positive, hospitalized, severe disease, and deceased subsets (**Table 1**). Multivariate logistic regression models were fit from the covariates against each of the outcome variables (hospitalization, severe disease, and death). The coefficients and standard error generated from this regression were converted to odds ratios, and p-values were generated using a Wald Test. The predictive capacity of each model was determined by re-fitting the model on 80% of the data, and then assessing its performance on the remaining 20%. The data was split such that the case to control ratio in each subset was equal. The true positive and false positive rates generated when comparing the predictions to true outcomes at a series of different prediction cut-offs were organized into a Receiver Operator Curve (ROC). The area under the ROC curve (AUC) was extracted to assess predictive ability. For all models, patients with unknown or missing data were included in the overall cohort to avoid bias, but the “Unknown” categories for each covariate was excluded from data visualization. R (version 4.0.1) was utilized for all computations.

## RESULTS

### Baseline Sample Clinical and Demographic Characteristics

Our patient cohort consists of 11,930 COVID-19 patients between March and August of 2020, which was 12% of patients tested in that period (**Supplemental Figure 1**). Nearly 50% of COVID-19 positive patients (n=4,895) were hospitalized in the NYP system in this timeframe. Of these hospitalized patients, 22% (n=1,070) developed severe disease, defined by intubation with vasopressor use, ARDS diagnosis, and/or kidney dialysis (**Supplemental Figure 1**). Deceased patients (n=1,654) were defined by the vital status in the EHR. **Supplemental Table 1** contains descriptive statistics of all sixteen variables (described in Methods) across the three outcome groups.

We utilized patients’ self-reported race and ethnicity identities, as described in methods. Our diverse multi-ethnic cohort resides across all five NYC boroughs (**Figure 1A)** and received COVID-19 tests at one of three sites in our network **(Supplemental Figure 3**). There is similar age distribution among race/ethnicity groups **(Figure 1B**), comparable to the general population (overall mean age = 57) (**Supplemental Figure 2**). More than 80% of racial/ethnic minority patients have moderate to high NDI (**Figure 1C and Supplemental Figure 4**), indicating they live in more deprived neighborhoods. Most patients reside in neighborhoods that are enriched for their same race (**Figure 1A**).

**Figure 1.**
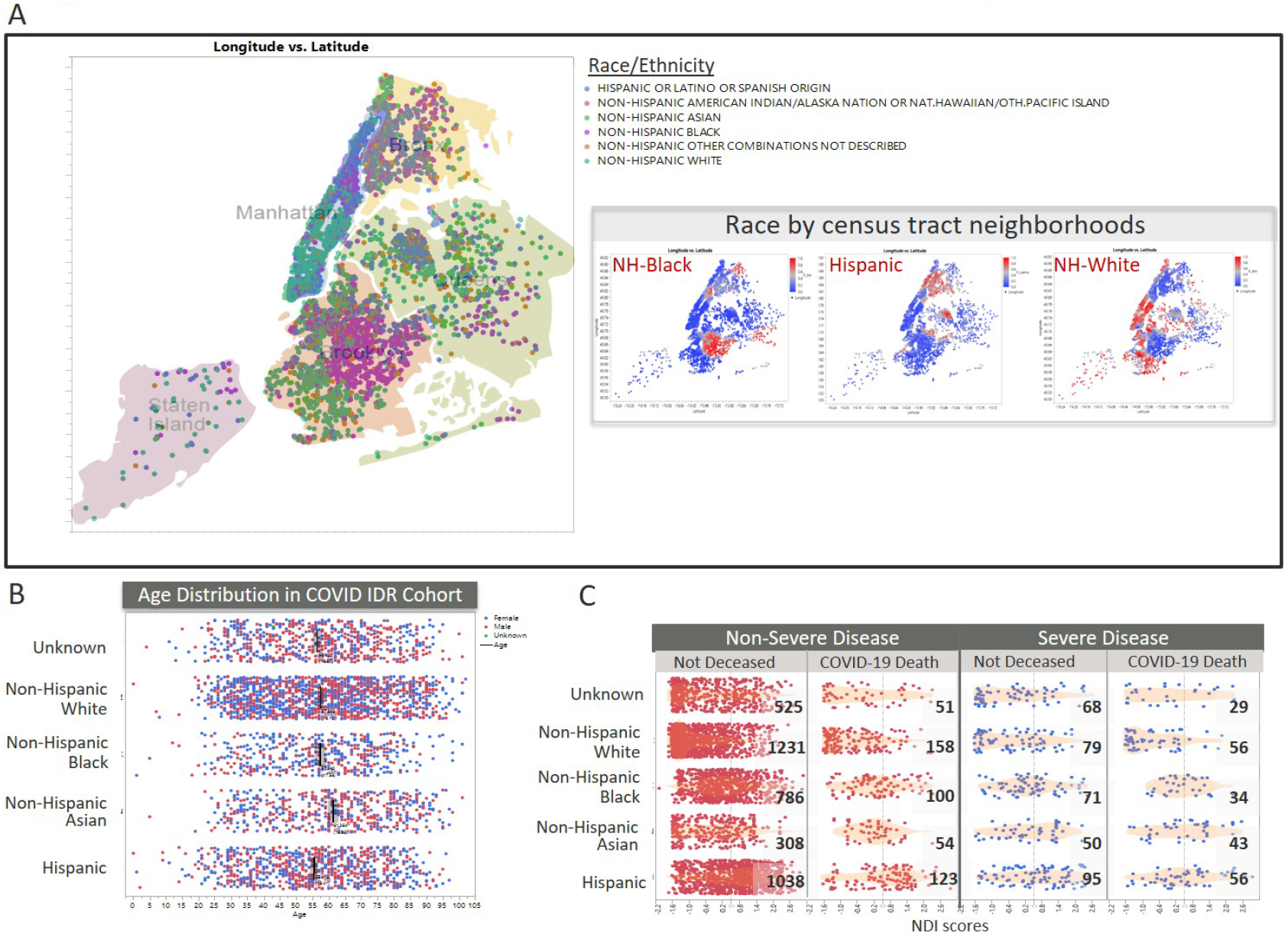
Residential area, NDI and race distribution of NYP COVID-19 patients are representative of general population. (A) Geographic distribution of COVD-19 patients by latitude-longitude coordinates, color-coded by self-reported race/ethnicity groups. Inset indicates the prevalence of specific race groups across the patient’s neighborhoods, based on census-track data. (B) Age distribution of cohort, by race. Bars indicate the mean age for each race group. Color coding of points indicate self-reported sex (Blue=Female, Red=Male). (C) Distribution of Neighborhood Deprivation Index (NDI) scores, by race. Column groups are based on our severe disease status model and mortality outcome models.

### Factors that impact risk of hospitalization among COVID-19 patients

We first determined factors associated with hospitalization of COVID-19 patients. Specifically, we quantified the risk of eighteen factors using a fully adjusted multivariate model (**Figure 2A)**. Similar to other studies^10^, we found risk of hospitalization increased with age, starting at age 30 (OR=1.79, p=7.91×10 ^-06^) up to 80+ (OR=5.78, p=2.29×10 ^-24^) and male sex (OR=1.50, p=5.92×10^−08^). Interestingly, among race/ethnicity variables, NH-Black and NH-Other patients had the strongest association in the hospitalization model, but for being less likely to be hospitalized (NH-B: OR=0.60, p=1.15×10^−04^; NH-O: OR=0.73, p=0.027. For clinical factors within the entire cohort, all co-variates were associated with increased odds of hospitalization except T1D, asthma, and obesity (HTN: OR=1.89, p=1.26×10^−10^; Depression: OR-1.76, p=3.07×10^−05^; T2D: OR=1.48, p=1.39×10^−04^; HF: OR=1.42, p=0.021; Cancer: OR=0.55, p=0.028; COPD: OR=1.49, p=0.045; CVD: OR=1.32, p=0.049). Lastly, there was an intriguingly high association with specific hospital testing sites, with Site 3 having the highest odds of hospitalization (OR=3.32, p=4.72×10^−22^). Insurance type was also shown to be significantly associated with hospitalization (Medicaid: OR=1.89, p=0.016; Medicare: OR=1.61, p=0.033). Overall, the clinical factors had the highest magnitude of risk, compared to other determinant types.

**Figure 2.**
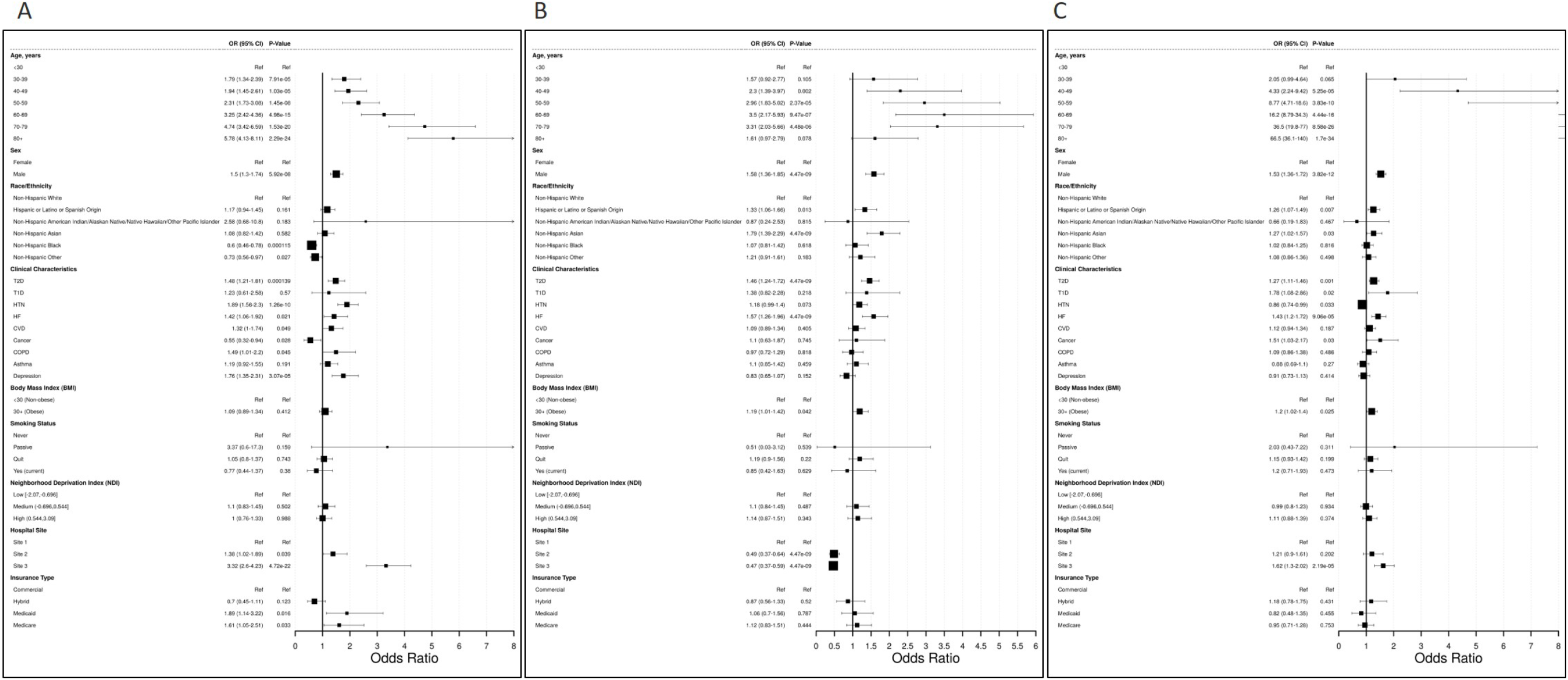
Fully adjusted multivariate models of hospitalization, severe disease, and mortality outcomes among confirmed COVID-19 patients. (A) Analysis of odds of hospitalization among all confirmed COVID-19 patients. (B) Analysis of odds of severe disease among hospitalization confirmed COVID-19 patients. (C) Analysis of odds of mortality among all confirmed COVID-19 patients. Analyses conducted using electronic health record data from NewYork-Presbyterian from March 1, 2020 to August 20, 2020. Odds Ratios shown with 95% confidence intervals in parentheses. NH = Non-Hispanic; T2D = Type 2 Diabetes; T1D = Type 1 Diabetes; HTN = Hypertension; HF = Heart Failure; CVD = Cardiovascular Disease; COPD = Chronic Obstructive Pulmonary Disease.

### Factors that impact risk of disease severity among COVID-19 patients

We then sought to determine the magnitude of risk for factors associated with severe disease among COVID-19 hospitalized patients, using a similar model approach as our hospitalization risk model (**Figure 2B**). Of the demographic factors, males were 1.58 times more likely to develop severe disease than females (p= 4.47×10^−09^). Risk of severe disease increased with age, starting at ages 40-49 (OR=2.30, p=0.002) up to ages 70-79 (OR: 3.31, p=4.48×10^−06^). NH-Asian and Hispanic patients were more likely to develop severe disease (NH-A: OR= 1.79, p= 4.47×10^−09^; HISP: OR=1.33, p=0.013), compared to NH-White patients. The most significant clinical factors associated with severe COVID-19 were type 2 diabetes (OR=1.46, p=4.47×10^−09^), heart failure (OR=1.57, p=4.47×10^−09^), and obesity (OR=1.19, p=0.042). Social determinants, such as NDI and insurance status had no significant impact on severe disease outcome overall. A significant association with specific hospital testing site was observed (Site 2: OR=0.49, p=4.47×10^−09^; Site 3: OR: 0.47, p=4.47×10^−09^). Thus, in this cohort, the sites associated with higher likelihood of hospitalization have a lower likelihood of developing severe disease after hospitalization.

### Factors that increase risk of mortality amongCOVID-19 patients

Lastly, we measured the impact of risk factors leading to death among confirmed COVID-19 patients, as opposed to recovering from infection (**Figure 2C**). Our third model includes 1,654 deceased patients whose death occurred following a COVID-19 diagnosis, without regard to hospitalization. Upon model fitting, we found that age was the most significant demographic factor associated with COVID-19 mortality, in a “dose-dependent” fashion, starting at ages 40-49 (OR=4.33, p=5.25×10^−05^) ranging to age 80+ (OR=66.5, p=1.70×10^−34^). Male sex (OR=1.53, p= 3.82×10^−12^), Hispanic ethnicity (OR=1.26, p=0.007), and NH-Asian race (OR=1.27, p=0.03) were also associated with mortality among COVID-19 patients. Type 2 diabetes (OR=1.27, p=0.001), type 1 diabetes (OR=1.78, p=0.02), hypertension (OR=0.86, p=0.033), heart failure (OR=1.43, p= 9.06×10^−05^), cancer (OR=1.51, p=0.033), and obesity (OR=1.20, p=0.025) were the all associated with odds of mortality. NDI, smoking status, and insurance type were not significant risk factors, after adjusting for the other variables.

### Differences in COVID-19 outcome risk factors across Patient Race/Ethnicity

Several reports have indicated the disproportionately higher rates of COVID-19 related hospitalization and mortality in the US Hispanic and NH-Black populations, compared to NH-White^1-6^. Our full cohort analyses have identified risk factors that are independent of race groups. To address whether some risk factors are more important in certain race groups, we fit new models within specific race groups, including NH-Black, NH-Asian, NH-White and Hispanic categories (**Figure 3**). Surprisingly, we found few risk factors that were consistent across race groups, with hypertension being the sole risk factor for hospitalization across all race groups (**Figure 3A. NH-W: OR=2.17, p=1.31×10**^**-04**^**; NH-B: OR=1.68, p=0.031; NH-A: OR=2.68, p=0.004; HISP: OR=2.09, p=3.44×10**^**-04**^). Age (NH-W: OR=3.17, p=4.87×10^−09^; NH-A: OR=3.42, p=7.97×10^−06^; HISP: OR=1.61, p=0.04) and depression (NH-W: OR=1.85, p=0.01; NH-A: OR=12, p=0.02; HISP: OR=1.93, p=0.018) were significant risk factors in all but the NH-Black population (**Figure 3A**). Male sex was associated with higher odds of hospitalization among NH-Asian patients (NH-A: OR=2.62, p=1.12×10^−04^) and Hispanic patients (HISP: OR=1.56, p=0.002). We also found several clinical risk factors for hospitalization were only significant in specific race groups. For example, type 2 diabetes was only a significant risk factor in NH-Black patients (NH-B: OR=1.81, p=0.014), heart failure was a significant risk factor in NH-Asian patients (NH-A: OR=4.31, p=0.037), COPD was a significant risk factor in NH-White patients (NH-W: OR=2.70, p=0.009). Interestingly, several variables that were not significantly associated with odds of hospitalization in the overall cohort model were identified as race-specific risk factors. These included cancer diagnoses in NH-Black and Hispanic patients (NH-B: OR=0.20, p= 0.005; HISP: OR=0.25, p= 0.022), smoking status (NH-A: OR_Quit_=0.23, p_Quit_=0.008; NH-A: OR_Yes (current or passive)_=0.02, p_Yes (current or passive_=0.006; HISP: OR_Yes (current or passive_=0.32, p_Yes (current or passive_=0.046), and insurance type (NH-W: OR_Hybrid_=0.41, p_Hybrid_=0.034; NH-B: OR_Hybrid_=3.11, p_Hybrid_=0.049, OR_Medicaid_=4.56, p_Medicaid_=0.028; HISP: OR_Medicare_=3.46, p_Medicare_=0.009) (**Figure 3B**). Lastly, NDI had differential impact across race groups (**Supplemental Figure 5**) where moderate NDI was associated with hospitalization in NH-Asian patients (NH-A: OR: 4.00, p=0.006) and high NDI scores were associated with hospitalization in NH-White patients (NH-W: OR=2.10, p=0.032).

**Figure 3.**
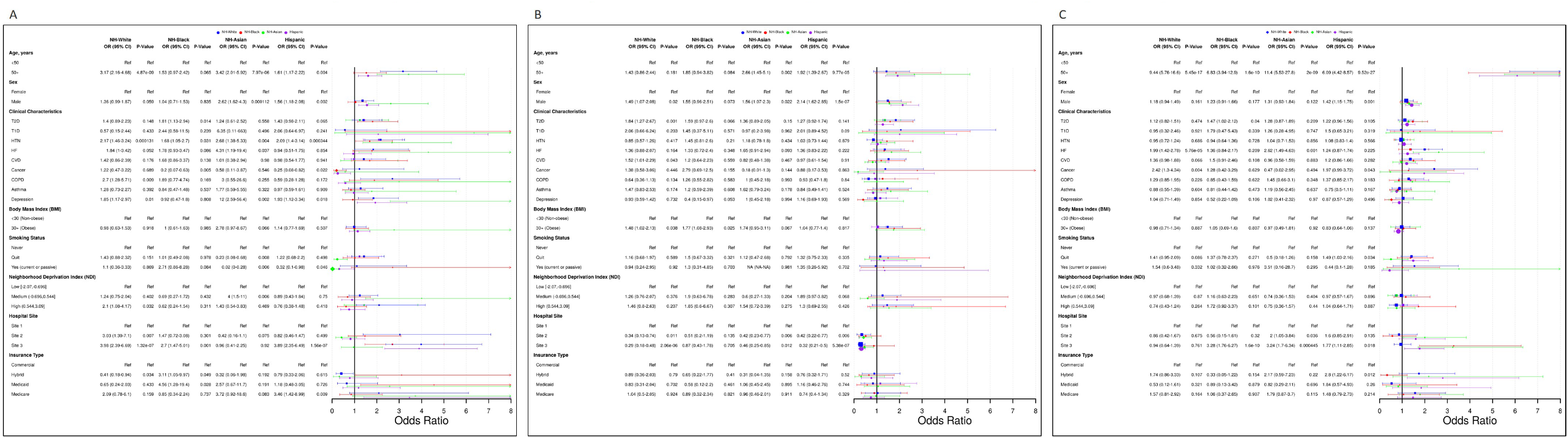
Fully adjusted multivariate models of hospitalization, severe disease, and mortality outcomes among confirmed COVID-19 patients, by race/ethnicity. (A) Odds of hospitalization among confirmed COVID-19, by race/ethnicity group. (B) Odds of severe disease among hospitalization confirmed COVID-19 patients, by race/ethnicity group. (C) Odds of mortality among all confirmed COVID-19 patients, by race/ethnicity group. Analyses conducted using electronic health record data from NewYork-Presbyterian from March 1, 2020 to August 20, 2020. Odds Ratios shown with 95% confidence intervals in parentheses. NH = Non-Hispanic; T2D = Type 2 Diabetes; T1D = Type 1 Diabetes; HTN = Hypertension; HF = Heart Failure; CVD = Cardiovascular Disease; COPD = Chronic Obstructive Pulmonary Disease.

For demographic variables in the race-group severe COVID-19 disease model (**Figure 3B**), surprisingly, age was only a significant severity risk factor among NH-Asian and Hispanic patients (NH-A: OR=2.66, p= 0.002; HISP: OR=1.92, p=9.77×10^−05^). Similarly, male sex was a significant severe disease risk factor for all but NH-Black patients, with the highest impact in Hispanic patients (NH-W: OR=1.49, p=0.02; NH-A: OR=1.56, p= 0.022; HISP: OR=2.14, p= 1.50×10^−07^). Across clinical factors, type 2 diabetes was a significant factor only in White patients (W: OR=1.84, p=0.001), cardiovascular disease was a race-specific risk factor of severe disease among NH-White patients (NH-W: OR=1.52, p=0.043), and obesity was a significant clinical risk factor among NH-White and NH-Black patients (NH-W: OR: 1.48, p=0.038; NH-B: OR=1.77, p=0.025). Odds of severe disease was lower among patients tested at hospital site 2 (NH-W: OR=0.34, p=0.011; NH-A: 0.42, p=0.006; HISP: OR=0.42, p=0.006) and site 3 (NH-W: OR=0.29, p=2.06×10^−06^; HISP: OR=0.32, p=5.38×10^−07^).

Finally, the only significant risk factors for increased odds of mortality among all race groups was age (NH-W: OR=9.44, p=5.45×10^−17^; NH-B: OR=6.83, p=1.60×10^−10^; NH-A: OR=11.40, p=2.00×10^−09^; HISP: OR=6.09, p=9.52×10^−27^) (**Figure 3C**). Male sex was associated with increased odds of mortality only among Hispanic patients (HISP: OR=1.42, p=0.01). Among clinical characteristics, only heart failure (NH-W: OR=1.99, p=5.76×10^−05^; NH-A: OR=2.62, p=0.001) and cancer (NH-W: OR=2.42, p=0.004; HISP: OR=1.97, p=0.043) were shown to be statistically significant risk factors of mortality within race groups. Among the Hispanic population, smoking status (HISP: OR_Quit_=1.49, p=0.034) and insurance type (HISP: OR_Hybrid_=2.80, p=0.012) were shown to be associated with increased odds of mortality. Surprisingly, NDI had relatively small impact in any race-group’s mortality risk, which suggests social determinants were not the primary drivers of mortality disparities, when considered in context with the risk associated with clinical factors.

### Predictive efficacy of the multivariate models

We hypothesized that the multivariate logistic regression models could be developed into predictive models. The predictive capacity of each model was assessed using the area under the receiver operator curve (AUC) as discussed in Methods. We found that all models had some predictive power; however, accuracy varied among outcomes (**Figure 4**). Overall, the hospitalization model was highly predictive in the overall cohort (AUC=0.969) (**Figure 4Ai**), while the severe disease and death models performed with lower accuracy (AUC=0.698 and AUC=0.812, respectively) (**Figure 4A-vi and xi**).

**Figure 4A.**
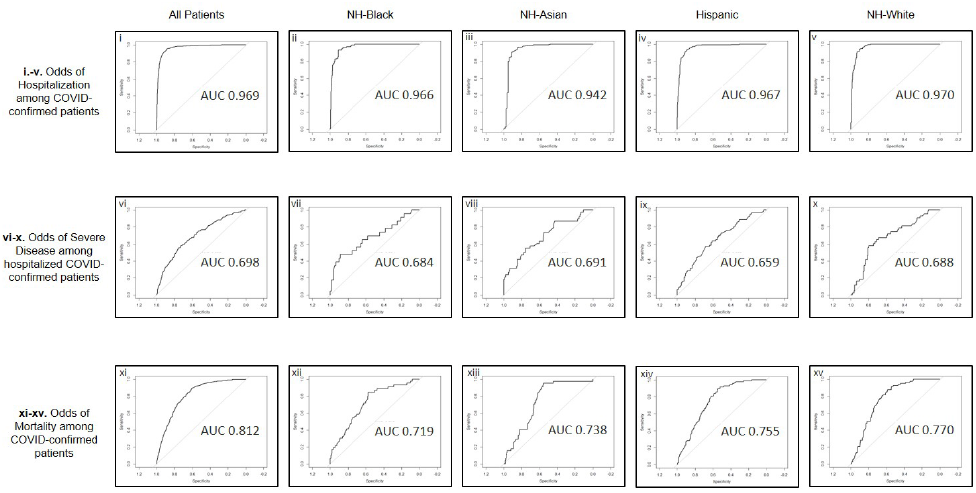
Performance of Risk Models and Ranked Risk Factors. **A**. Receiver operating characteristic (ROC) curves and areas under the curves (AUC) for each fully adjusted multivariate regression model: (i.-v) odds of hospitalization among confirmed COVID-19 patients overall and by race groups; (vi-x) odds of severe disease among hospitalized confirmed COVID-19 patients overall and by race groups; (xi-xv) odds of mortality among confirmed COVID-19 patients overall and by race groups. NH = Non-Hispanic.

Using models fitted with only specific race groups, we found that accuracy of the hospitalization models remains high across all 4 racial/ethnic groups, with NH-White group having the highest accuracy (NH-W: AUC=0.970, **Figure 4A-v**) and the NH-Asian group having the lowest accuracy (NH-A: AUC=0.942, **Figure 4A-iii**). Severe disease risk models were least accurate overall, ranging from the lowest AUC in the Hispanic group (HISP: AUC=0.659, **Figure 4A-ix)** to the highest AUC among the NH-Asian group (NH-A: AUC=0.691, **Figure 4A-viii**). The death risk models performed with high accuracy, but to varying degrees across race groups. The NH-Black group had the lowest accuracy with our model (AUC=0.719, **Figure 4A-xii**), while NH-White patients had the highest accuracy (AUC=0.770, **Figure 4A-xv**). The attenuated accuracy of our models in specific race groups likely reflects the differential influence of each factor toward disease outcomes among race groups.

## DISCUSSION

Overall, our findings in this large New York City cohort (n=11,930) of COVID-19 patients, diagnosed between March and August of 2020, present a unique perspective of relative risks impacting disparities. While we confirm the vulnerabilities of certain race/ethnicity groups, we also uncovered that typical demographic and social determinants that were implicated in COVID-19 disparities were not as impactful as the clinical factors. Interestingly, we also observed differences in which factors associated with severe disease risk of hospitalized patients among race-groups.

Several of our results agree with recently published reports from other academic health centers^10^ and the Veteran Affairs Hospitals^6,11^, including the impact of age and sex on hospitalization and severe outcomes. One consistently surprising result among these studies was that NH-Black patients were less likely to be hospitalized after a COVID-19 diagnosis, compared to other race groups. This is despite public health data reports indicating disproportionately high rates of NH-Black hospitalization, nation-wide. There may be several reasons for this difference. First, NH-Black patients in our cohort, like other large academic institutions, may represent a specific subset of the greater NYC NH-Black population, which illustrates the jeopardy in generalizations of race-group risk when these populations are distributed across broad social strata. Secondly, based on their residential locations, NH-Black patients in our cohort were likely to travel beyond their boroughs for access to our hospital testing sites (**Supplemental Figure 3**). At the peak of COVID-19, this was likely necessary because of the limited access to testing in certain areas. This also presents a possibility that, if their COVID-19 condition escalated to require urgent care or hospitalization, they may have sought care in alternative healthcare systems in closer proximity to their residence, which we would not be able to track in our cohort data sources.

Our study describes race-group specific risk factors, compared to other publications that generalize risk factors to be similar across the entire population at-large. For instance, in our hospitalized severe disease model, we found that age was only significant in the NH-Asian and Hispanic groups, while obesity was the only significant factor for severe disease in NH-Black patients. This suggests that generalized use of risk factors for prioritization models across the population at-large would inherently benefit certain race/ethnicity groups more than others, de-prioritizing the high-risk status of specific race groups, leading to greater disparities across the continuum of disease management and prevention.

The results of this study can have immediate transformative clinical impact. Our use of fully adjusted multivariate models assesses probability of severe outcomes in the mixed context of social, demographic, and clinical factors, providing a comprehensive approach to reduce racial disparities through prioritization of risk. Ranked severe disease risk factors, by effect size across all models (**Figure 4B**), gives insight to differential disease course as well as a potential plan of prioritization. First, the impact of Type 2 Diabetes on severe disease was significant in NH-White patients, but not in NH-Black patients after adjusting for BMI. This suggests that while this disorder is typically more prevalent in the NH-Black population, obesity can influence its role in COVID-19 outcomes. The differential impact of clinical risk factors across race groups may indicate population-level differences in chronic disease management/status may influence physiology and biological determinants of COVID-19 disease trajectory. Next, we found that hospitalization is a benefit to outcome. Specifically, in the hospital sites where hospitalization was more likely (**Figure 2A**), severe disease was less likely (**Figure 2B**). This may suggest that severe disease can be avoided, for patients treated at certain sites, if hospitalization occurs.

**Figure 4B.**
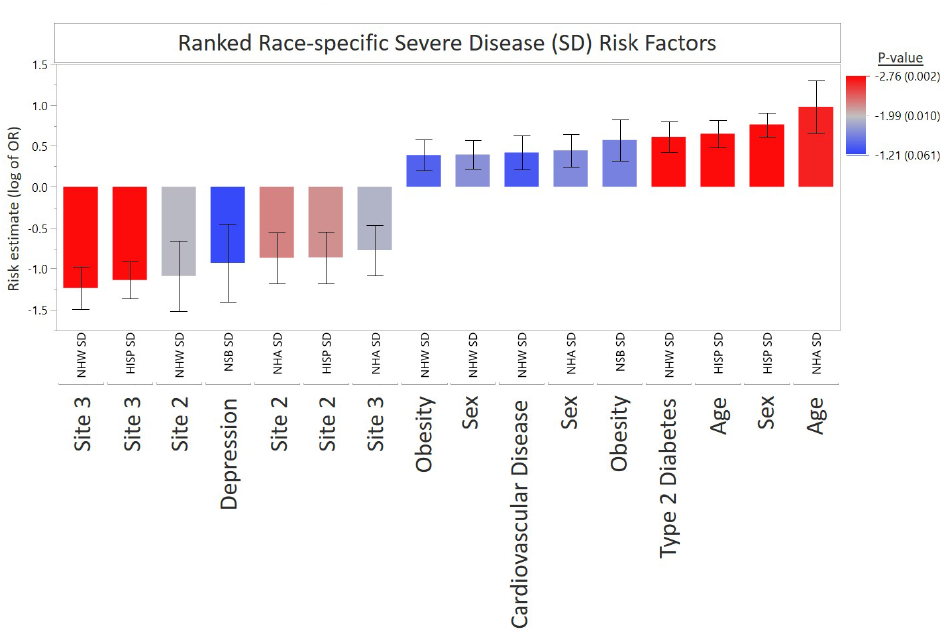
Ranked risk factors in severe disease models. Combined significant severe disease risk factors identified in the all-inclusive models and stratified within race groups. Bar graphs represent estimated effect measurements for indicated subset and factors, with Standard Error bars. P-values are color coded with increasing significance, red=most significant, blue=least significant.

This finding was consistent for most race groups, excluding NH-Black patients (**Figures 3B and 4B**) who were also found to be less likely to be hospitalized.

There are important limitations to consider in interpreting our results. First, our study has the typical limitations RWD data collection from EHR systems. However, use of RWD is advantageous because in the acute and fast-paced context of a deadly pandemic, urgency of saving lives precedes the priority of appropriate study designs and specialized data collection. However, completeness and accuracy of some variables that are typically part of self-reported intake surveys, including smoking status, medical history and loss of follow-up is debatable. The NDI application of SDOH is a powerful indicator of patients’ ‘placemats’ or integrated neighborhood-level determinants, which is often more predictive of health outcomes than direct measures of socioeconomic status. However, the NDI does not directly measure some key factors, such as systemic racism or social capital^12^ though the measures included in calculating the NDI are certainly influenced by these factors. (**Supplemental Figure 3**).

## CONCLUSIONS

In conclusion, our large cohort study of over 11,000 COVID-19 patients reveals a slightly different perspective than what has previously been reported nationwide. We find that clinical comorbidity factors, such as Type 2 Diabetes, are more significant to COVID-19 outcome than social determinants. We also found risk factors are not generalizable across all race groups. Finally, we demonstrate that poor outcomes such as hospitalization risk can be predicted reliably using pre-infection data. This indicates that individuals at risk can in theory be reliably identified and triaged for highly effective care including vaccination.

## Data Availability

The data included in this study are the direct output of the hospital medical record system and can not be shared in its raw form. A summary table is included. Summary data can be supplied upon request to the Weill Cornell Informatics Department.

## Acknowledgements

This work was funded by several awards to the co-authors; including a COVID-19 Research Grant from the Weill Cornell Office of Research (MBD), a NIH (National Institutes of Health) COVID-19 Disparities Supplement for the WCM CTSC (Clinical and Translational Science Center) NIH UL1TR002384 grant (JIM, OE, SI, MBD, JM).

## TABLES

**Supplemental Table 1.**
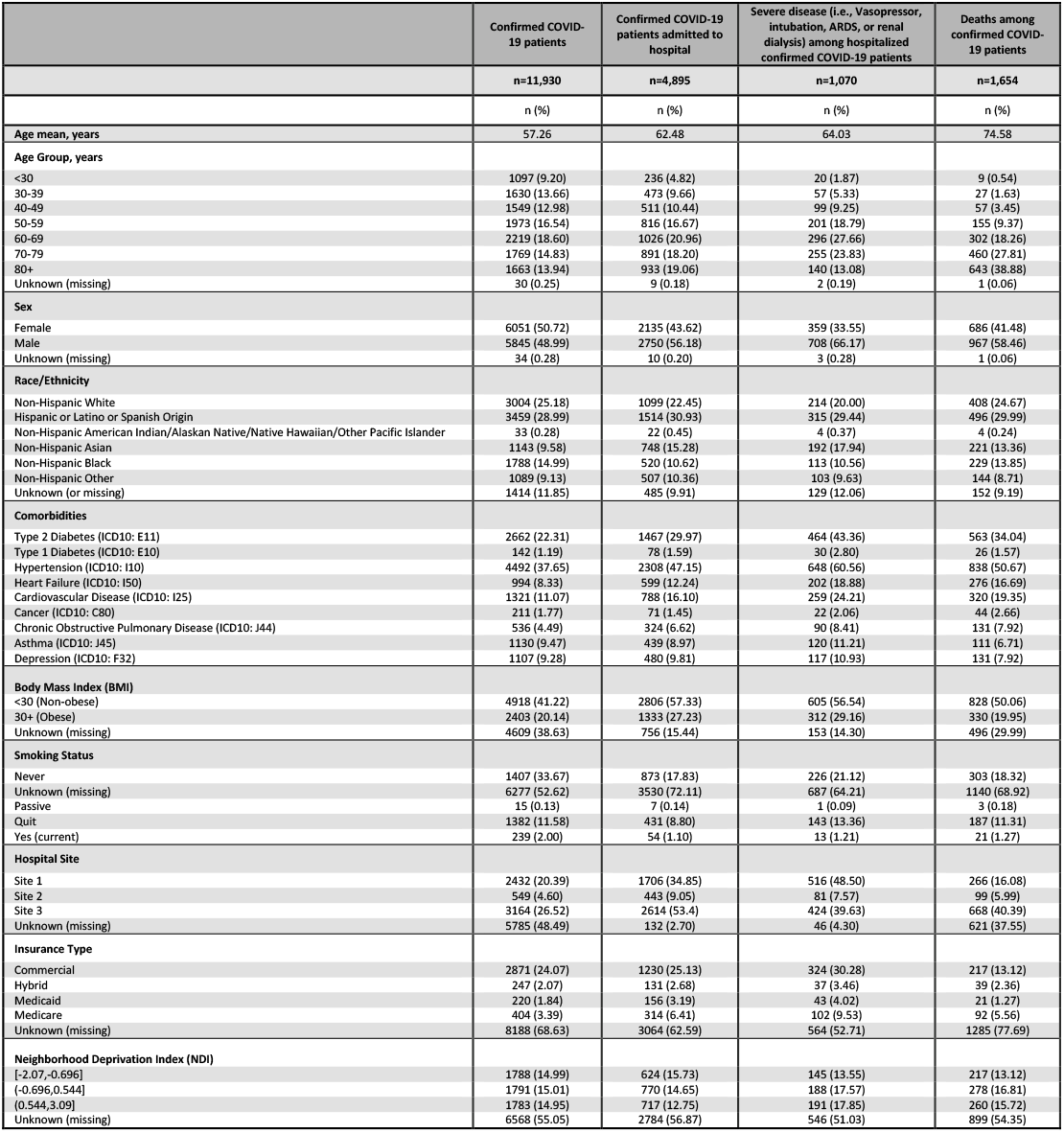
Descriptive statistics of demographic, clinical, and socioeconomic factors of confirmed COVID-19 patients. Analysis of electronic health record data from NewYork-Presbyterian from March 1, 2020 to August 20, 2020.

## FIGURE TITLES AND LEGENDS

**Supplemental Figure 1. Study Flow Chart**. Patient population in New York Presbyterian COVID-19 Data repository. NYP = NewYork-Presbyterian; IDR = Institutional Data Repository; EHR = Electronic Health Record.

Supplemental Figure 2. Age distribution across the full dataset and individual race groups are comparable to the NYC demographics. (A) Histogram of the age groupings in the COVID IDR indicate that ∼60% of our population falls within the age range of 20-65. (B) This corresponds to the 2019 NYC census where 58% of the population is within the age range of 18-65 (panel B). (C) Age distribution across each race group shows correlated average age around 55-57, except for NH-Asians having a slightly higher average age. (D) COVID IDR age groups by decade show the generally even spread of age, indicating little to no bias in our dataset.

**Supplemental Figure 3. Testing sites compared to residential locations of patients**. Residential map of patients, color coded for the testing site where they received a COVID-19 test result. Most of the patients were tested in the vicinity of their residents, though a significant proportion tested in different boroughs than where they resided. The majority of Brooklyn residents tested at Site 1.

**Supplemental Figure 4. Box plots of NDI distribution by race/ethnicity for primary outcomes of interest:** (A) Confirmed COVID-19 patients; (B) Hospitalized confirmed COVID-19 patients; (C) Severe disease among hospitalized confirmed COVID-19 patients; (D) Deceased confirmed COVID-19 patients. NH = Non-Hispanic.

**Supplemental Figure 5. Distribution of NDI scores for patient race groups by hospital testing sites**. Boxplots indicate the NDI range and average for each race group that utilized the indicated hospital sites. Floating blue bar indicates hospital group NDI score average. This indicates that each hospital has a range of patients from areas with varying NDI. Overall, Site 2 had the highest NDI and Site 1 had the lowest average NDI. The contrast of white patients at Site 2 indicates that NDI can vary greatly between race groups in the same region of a borough, a characteristic that reflects red-lining and systemic racial bias.

## References

1. Argenziano MG, Bruce SL, Slater CL, et al. Characterization and clinical course of 1000 patients with coronavirus disease 2019 in New York: retrospective case series. BMJ. 2020;369:m1996.

2. Hanson AE, Hains DS, Schwaderer AL, Starr MC. Variation in COVID-19 Diagnosis by Zip Code and Race and Ethnicity in Indiana. Front Public Health. 2020;8:593861.

3. Chen JT, Krieger N. Revealing the Unequal Burden of COVID-19 by Income, Race/Ethnicity, and Household Crowding: US County Versus Zip Code Analyses. J Public Health Manag Pract. 2021;27 Suppl 1, COVID-19 and Public Health: Looking Back, Moving Forward:S43-S56.

4. Munoz-Price LS, Nattinger AB, Rivera F, et al. Racial Disparities in Incidence and Outcomes Among Patients With COVID-19. JAMA Netw Open. 2020;3(9):e2021892.

5. Little C, Alsen M, Barlow J, et al. The Impact of Socioeconomic Status on the Clinical Outcomes of COVID- 19; a Retrospective Cohort Study. J Community Health. 2021.

6. Kabarriti R, Brodin NP, Maron MI, et al. Association of Race and Ethnicity With Comorbidities and Survival Among Patients With COVID-19 at an Urban Medical Center in New York. JAMA Netw Open. 2020;3(9):e2019795.

7. Sholle ET, Kabariti J, Johnson SB, et al. Secondary Use of Patients’ Electronic Records (SUPER): An Approach for Meeting Specific Data Needs of Clinical and Translational Researchers. AMIA Annu Symp Proc. 2017;2017:1581–1588.

8. Messer LC, Laraia BA, Kaufman JS, et al. The development of a standardized neighborhood deprivation index. J Urban Health. 2006;83(6):1041–1062.

9. Blueprint WHORD. Novel Coronavirus: COVID-19 Therapeutic Trial Synopsis. World Health Organization February 2020 2020.

10. Ogedegbe G, Ravenell J, Adhikari S, et al. Assessment of Racial/Ethnic Disparities in Hospitalization and Mortality in Patients With COVID-19 in New York City. JAMA Netw Open. 2020;3(12):e2026881.

