## Supplemental figures for "Social and Clinical Determinants of COVID-19 Outcomes: Modeling Real-World Data from a Pandemic Epicenter"

eFigure 1. Study Flow Chart.

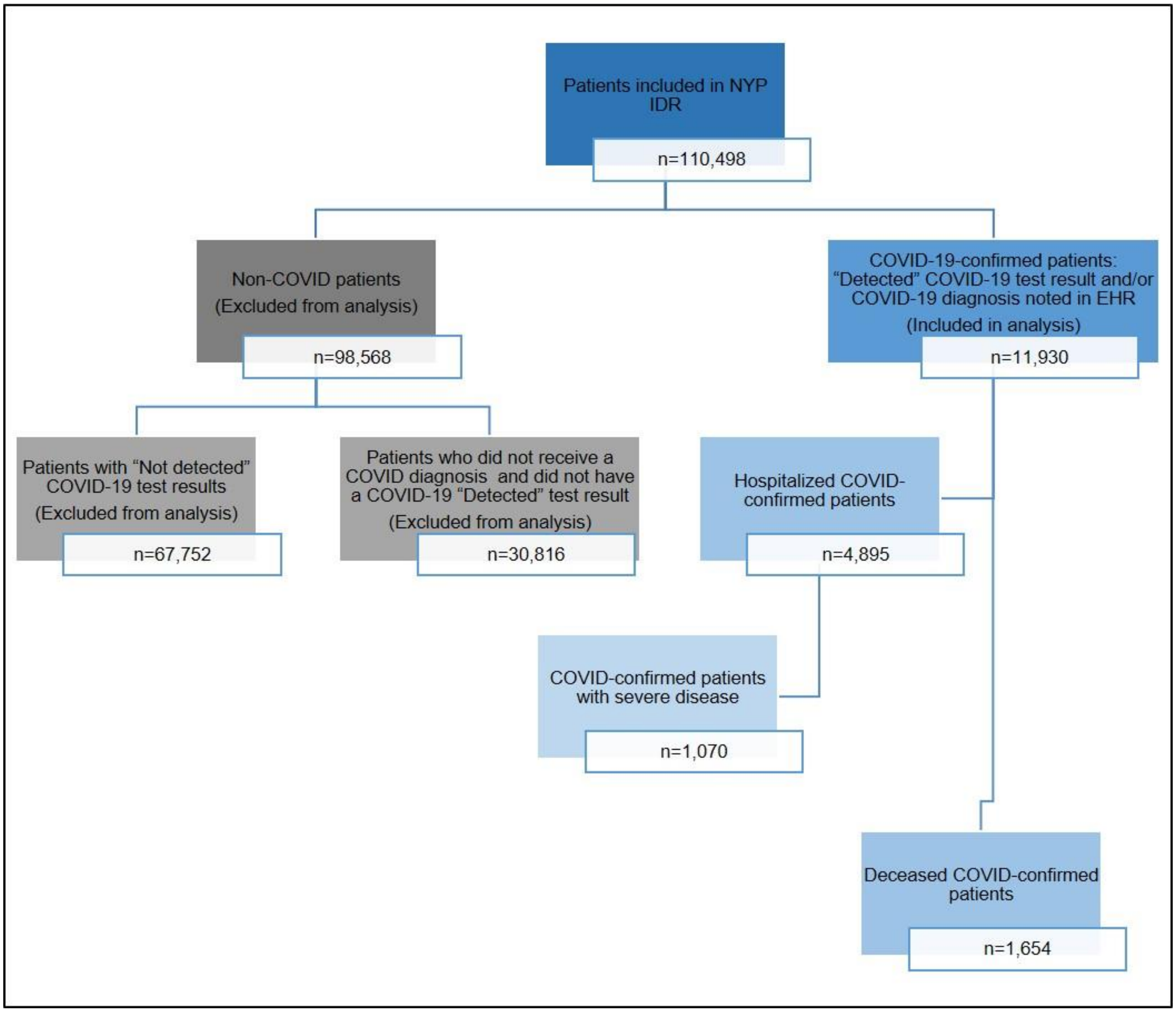


**eFigure 1. Study Flow Chart.** Patient population in New York Presbyterian COVID-19 Data repository. NYP = NewYork-Presbyterian; IDR = Institutional Data Repository; EHR = Electronic Health Record.

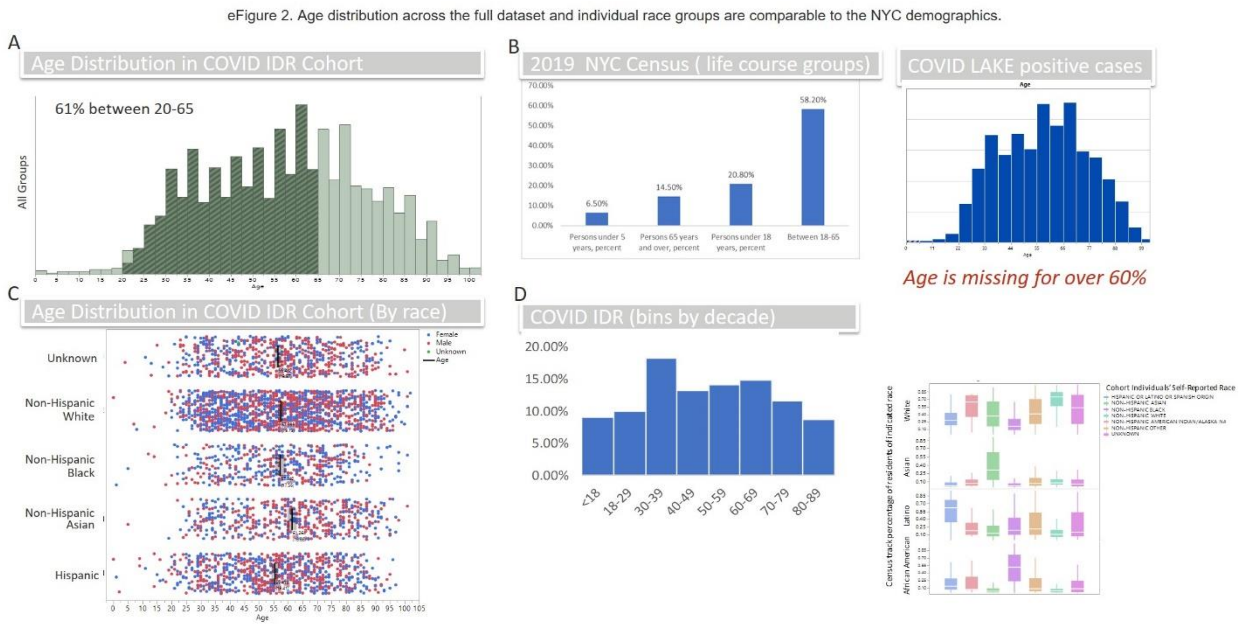


**eFigure 2. Age distribution across the full dataset and individual race groups are comparable to the NYC demographics.** (A) Histogram of the age groupings in the COVID IDR indicate that ~60% of our population falls within the age range of 20-65. (B) This corresponds to the 2019 NYC census where 58% of the population is within the age range of 18-65 (panel B). (C) Age distribution across each race group shows correlated average age around 55-57, except for NH-Asians having a slightly higher average age. (D) COVID IDR age groups by decade show the generally even spread of age, indicating little to no bias in our dataset.

eFigure 3. Testing sites compared to residential locations of patients.

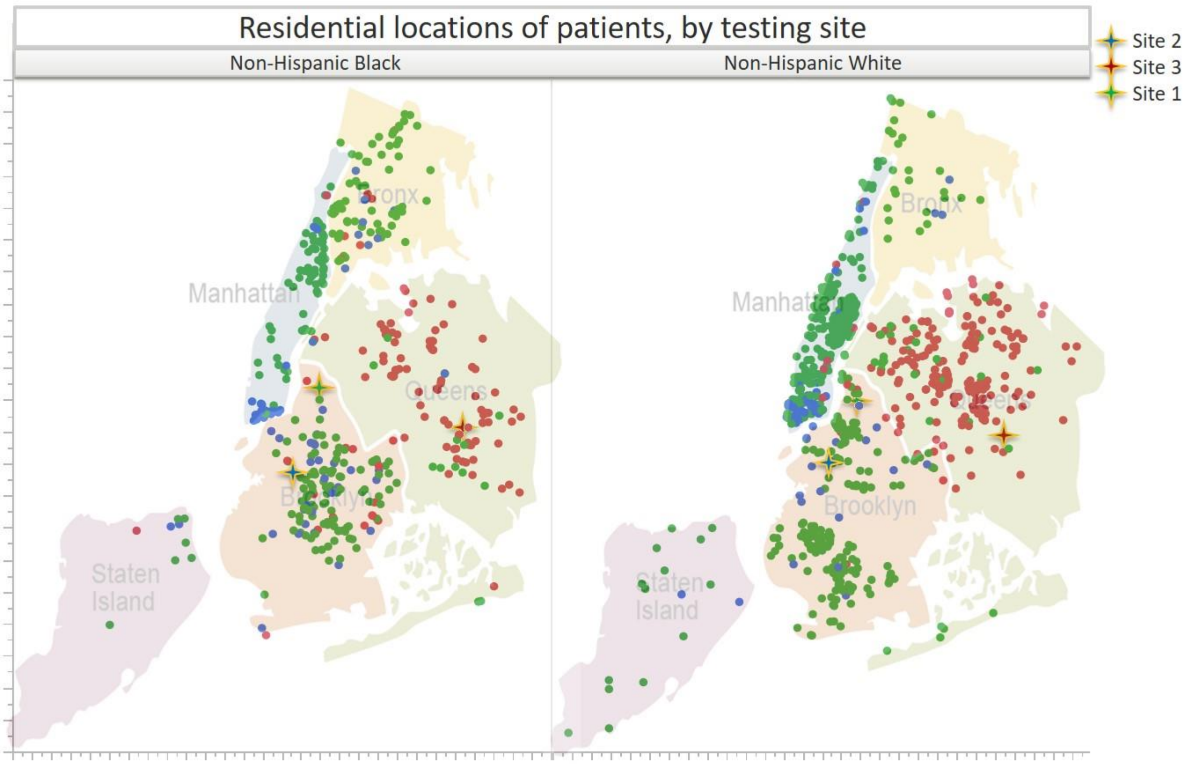


**eFigure 3. Testing sites compared to residential locations of patients.** Residential map of patients, color coded for the testing site where they received a COVID-19 test result. Most of the patients were tested in the vicinity of their residents, though a significant proportion tested in different boroughs than where they resided. The majority of Brooklyn residents tested at Site 1.

eFigure 4. Box plots of NDI distribution by race/ethnicity for primary outcomes of interest.

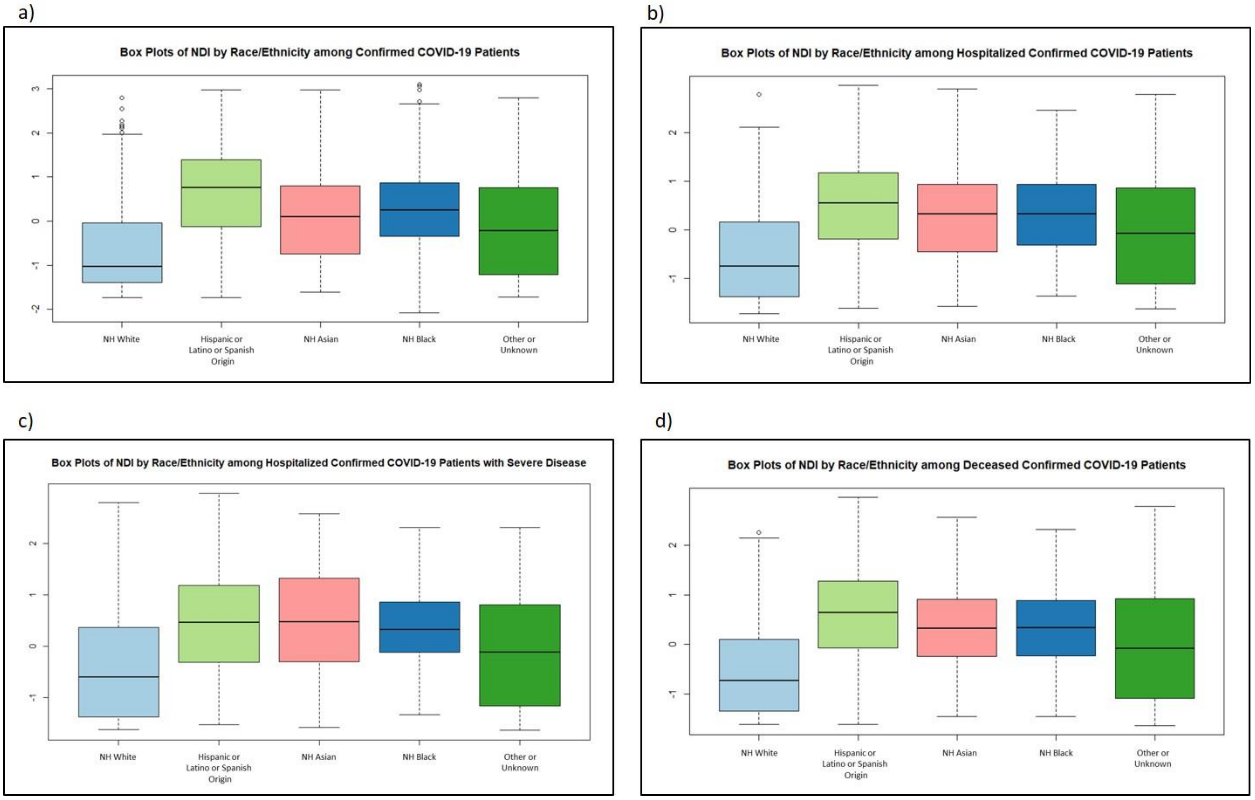


**eFigure 4. Box plots of NDI distribution by race/ethnicity for primary outcomes of interest:** (A) Confirmed COVID-19 patients; (B) Hospitalized confirmed COVID-19 patients; (C) Severe disease among hospitalized confirmed COVID-19 patients; (D) Deceased confirmed COVID-19 patients. NH = Non-Hispanic.

eFigure 5. Distribution of NDI scores for patient race groups by hospital testing sites

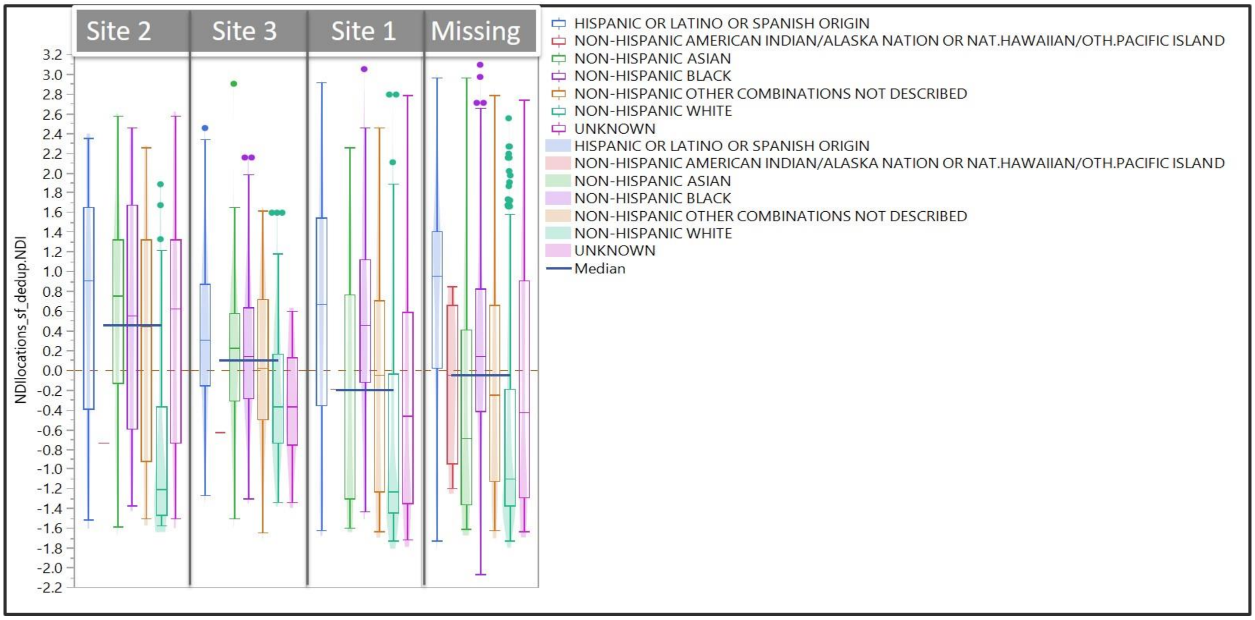


**eFigure 5. Distribution of NDI scores for patient race groups by hospital testing sites.** Boxplots indicate the NDI range and average for each race group that utilized the indicated hospital sites. Floating blue bar indicates hospital group NDI score average. This indicates that each hospital has a range of patients from areas with varying NDI. Overall, Site 2 had the highest NDI and Site 1 had the lowest average NDI. The contrast of white patients at Site 2 indicates that NDI can vary greatly between race groups in the same region of a borough, a characteristic that reflects red-lining and systemic racial bias.
